# An Ensembled Deep Learning Model Outperforms Human Experts in Diagnosing Biliary Atresia from Sonographic Gallbladder Images

**DOI:** 10.1101/2020.06.09.20126656

**Authors:** Wenying Zhou, Yang Yang, Cheng Yu, Juxian Liu, Xingxing Duan, Zongjie Weng, Dan Chen, Qianhong Liang, Fang Qing, Jiaojiao Zhou, Hao Ju, Zhenhua Luo, Weihao Guo, Xiaoyan Ma, Xiaoyan Xie, Ruixuan Wang, Luyao Zhou

## Abstract

It is still difficult to make accurate diagnosis of biliary atresia (BA) by sonographic gallbladder images particularly in rural area lacking relevant expertise. To provide an artificial intelligence solution to help diagnose BA based on sonographic gallbladder images, an ensembled deep learning model was developed based on a small set of sonographic images. The model yielded a patient-level sensitivity 93.1% and specificity 93.9% (with AUROC 0.956) on the multi-center external validation dataset, superior to that of human experts. With the help of the model, the performance of human experts with various levels would be improved further. Moreover, the diagnosis based on smartphone photos of sonographic gallbladder images through a smartphone app and based on video sequences by the model still yielded expert-level performance. Our study provides a deep learning solution to help radiologists improve BA diagnosis in various clinical application scenarios, particularly in rural and undeveloped regions with limited expertise.

## Introduction

Biliary atresia (BA) is a rare disease of infancy that affects varying lengths of both intrahepatic and extrahepatic bile ducts ^1^, about 1 in 5000-19000 infants all over the world ^2, 3, 4, 5, 6^. It is the most common cause for liver transplantation in infants younger than one year old ^7^. Optimal clinical outcome often needs timely diagnosis and Kasai portoenterostomy (KPE) surgery before two months old, which is associated with longer native liver survival ^8, 9, 10^. However, early identifying BA remains challenging in infants with cholestasis. Researchers have endeavored to screen the direct bilirubin concentration ^11^ or stool color ^4, 10^ in newborns and infants for early identification of BA and showed good results (with sensitivities of 97.1% ∼100%). Recently, serum matrix metalloproteinase-7 was reported as an effective diagnostic biomarker for BA, with sensitivity of 94%∼98.7% ^12, 13^. However, these tests are high resource-consumed and might be impractical in many countries and areas with underdeveloped healthcare conditions.

Ultrasound examination, due to its radiation-free and low-cost noninvasive property, is still the most-widely used method for initial detection of BA in jaundiced infants particularly in developing Asian countries like China and India ^14, 15, 16, 17^. Gallbladder abnormality is one of the most popular sonographic features used to identify BA ^18, 19, 20, 21^. As previously reported, gallbladder abnormalities can yield both sensitivities and specificities higher than 90% in experienced hand for the diagnosis of BA ^22^. However, it is still difficult to make a correct diagnosis by ultrasound examination mainly due to the lack of expertise in both diagnosis and management of BA in most hospitals particularly located in underdeveloped regions. Consequently, a substantial proportion of potential BA patients are often misdiagnosed followed by inappropriate treatments, and the average age of BA patients at KPE surgery was delayed to older than 70 days in China ^23^.

To improve the accuracy of BA diagnosis especially for underdeveloped countries or regions, one potentially promising way is to make use of the artificial intelligence (AI) techniques. Among the AI techniques, deep learning models, particularly the convolutional neural networks (CNNs), have been shown superior or comparable to human experts in many medical data analysis tasks, such as the diagnosis of skin cancers, localization and identification of polyps, and lung cancer screening ^24, 25, 26, 27, 28, 29^. However, as far as we know, no AI model has been developed for diagnosis of BA, partially because it is very time-consuming to collect enough number of medical data for BA, while deep learning models are often built on large-scale data. Considering the fact that ultrasound examination is very common in both primary and tertiary hospitals in China, any well-developed AI model based on sonographic gallbladder images for diagnosis of BA would alleviate the shortage of expertise in primary hospitals and may largely improve the diagnosis accuracy for the rare disease.

The purpose of this study is to develop a ensembled deep learning model(EDLM) for automatically and accurately identifying BA in infants with conjugated hyperbilirubinemia, based on limited number of sonographic gallbladder images collected from multicenter, and to help doctors improve their diagnosis of BA.

## Results

### Internal evaluation of the ensemble deep learning approach

The ensemble deep learning approach was firstly evaluated in a 5-fold cross validation manner on the training cohort. Specifically, the training cohort was partitioned into five complementary subsets of an equivalent number of patients. Then, every time four of the subsets were used as a training dataset to train an ensembled deep learning model, and the ensembled model was then applied to predict the category of each image in the remaining one (testing) subset. Such a process was repeated five times, each time using a unique subset as the testing dataset.

At both the image level and the patient level, the EDLMoutperformed the two experts in diagnosing BA, with the image-level sensitivity 88.2%, specificity 89.8%, and accuracy 89.4% of the model versus the sensitivity 93.8%, specificity 53.7%, and accuracy 63.7% of the most experienced expert, and the patient-level sensitivity 93.3% and specificity 85.2% of the model versus the sensitivity 90% and specificity 57.6% of the most experienced expert (Table 1). The ROC curves of the model at both levels also confirmed its superior performance over human experts (Figure 1).

**Table 1.** The diagnostic performance of the ensembled deep learning model and two human experts on the internal cross-validation dataset.

|  |  | AUROC | Sensitivity<br>(%) | Specificity<br>(%) | Accuracy<br>(%) | PPV (%) | NPV (%) |
| --- | --- | --- | --- | --- | --- | --- | --- |
| Image-level | AI Model | 0.952 | 88.2 | 89.8 | 89.4 | 74.1 | 95.8 |
|  | Expert A | NA | 76.3 | 91.0 | 87.4 | 73.9 | 92.0 |
|  | Expert B | NA | 93.8 | 53.7 | 63.7 | 40.3 | 96.3 |
| Patient-level | AI Model | 0.953 | 93.3 | 85.2 | 87.6 | 72.0 | 96.9 |
|  | Expert A | NA | 65.8 | 96.8 | 87.8 | 89.3 | 87.4 |
|  | Expert B | NA | 90.0 | 57.6 | 67.0 | 46.8 | 93.4 |

Note\_PPV= Positive predictive value; NPV= Negative predictive value.

**Fig. 1.**
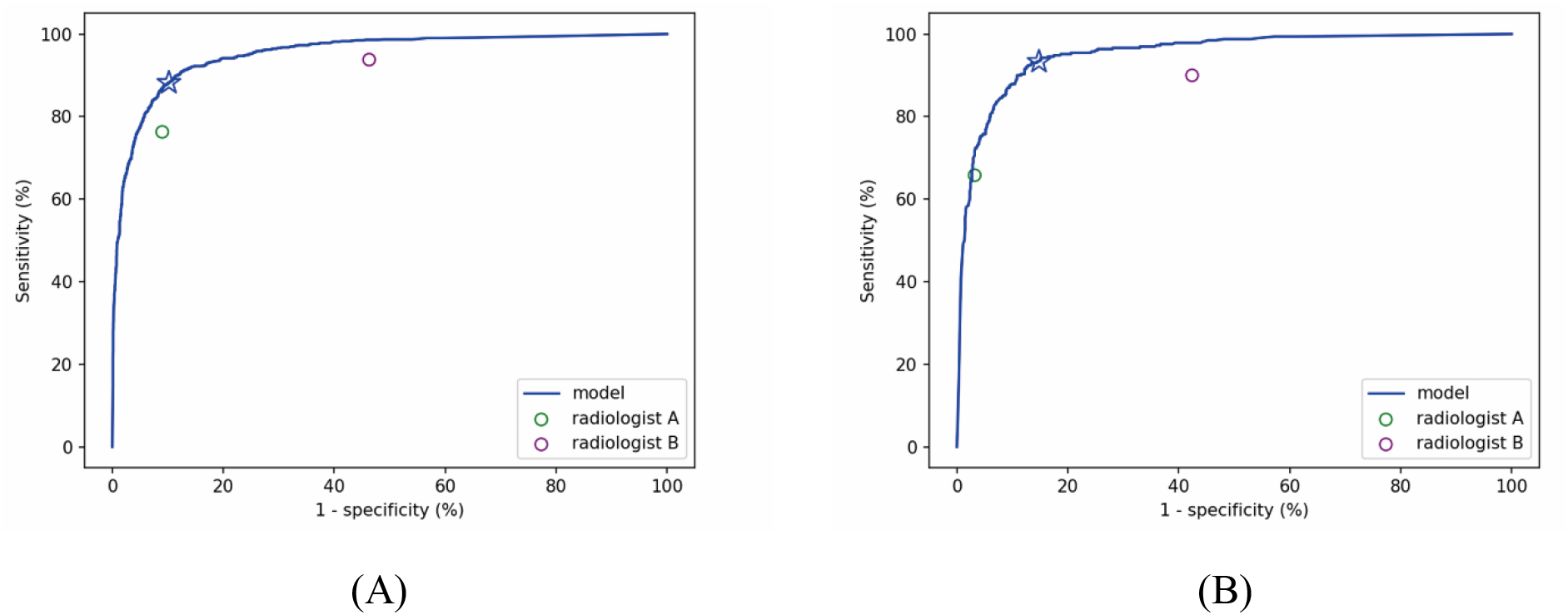
The ROC curves of the ensembled deep learning models for the diagnosis of biliary atresia on the internal cross-validation dataset with two human experts’ performance for comparison. (A) The ROC curve of the model at image level. (B) The ROC curve of the model at patient level. The performance of the two experts were represented by individual solid circle, which is under the ROC curve and therefore suggests inferior performance to the ensembled deep learning model. The blue star mark represents the performance of the model with the default threshold (0.5) to binarize predictions of the model.

### Robustness of the AI models to various scanning conditions

Considering that the trained deep learning model could be employed to various hospitals in which ultrasound scanning conditions may be different from that of the data for model training, the ensemble deep learning approach was also evaluated in terms of its robustness to screening machines, transducer frequencies and scanning period.

In the training cohort, images were retrospectively divided into three subsets based on whether the images were obtained from machines of brand Mindray, Supersonic, or the others (including TOSHIBA, Siemens, Samsung, HITACHI, ALOKA, Philips, GE and Esaote); or into two subsets based on whether the images were obtained by transducers of frequencies ≥14MHz or by transducers of frequencies <14MHz; or into two subsets based on whether the images were obtained before year 2018 or thereafter. For each scanning factor, with every unique subset of images as the validation dataset and the remaining subset(s) as the training dataset, the sensitivity of the trained EDLM was roughly in between those of two human experts (supplementary Tables S1, S2 and supplementary Figures S1, S2), supporting that the ensembled deep learning models were robust enough to be employable to different medical centers and for different screening machines. Furthermore, when using images of mediocre quality (with frequency<14MHz, scanning period ≤2018, supersonic + others or Mindary + others) to train the model and using the remaining subset(s) of good quality for validation, we found that the diagnostic performance of the model was higher than that when the training set and the testing set were reversed (AUROC 0.931 versus 0.835 for transducer frequency, 0.900 versus 0.832 for screening time, 0.950 and 0.807 versus 0.787 for screening machine, respectively at image level) (supplementary Table S1), which indicated that the quality of the image used for intelligent diagnosis should be as good as possible.

### External validation of the EDLM

More strictly, the effectiveness of the EDLM was evaluated by external validation with ultrasound images obtained from the other six hospitals. The EDLM yielded an image-level accuracy 92.3%, sensitivity 88.6%, specificity 93.7%, positive predictive value 84.6%, and negative predictive value 94.5%, respectively, clearly outperforming the three experts whose diagnosis sensitivities are 69.5%,77.1% and 87.3%, and specificities are 90.2%, 83.5% and 90.2%, respectively (Table 2, rows 1 to 4). The superior performance of the model can be also seen from the ROC curve of the EDLM(Figure 2A). Specially, 12 images of BA were misdiagnosed as non-BA by all three experts but were accurately identified by the EDLM.

**Table 2.** The diagnostic performance of the ensembled deep learning model, three experts, and the human-model combination on the external validation dataset and the comparison of the model and three experts based on sonographic videos.

|  |  | AUROC | Sensitivity<br>(%) | Specificity<br>(%) | Accuracy<br>(%) | PPV<br>(%) | NPV<br>(%) |
| --- | --- | --- | --- | --- | --- | --- | --- |
| Image-level | AI Model | 0.942 | 88.6 | 93.7 | 92.3 | 84.6 | 94.5 |
|  | C | NA | 77.1 | 83.5 | 91.7 | 64.5 | 90.3 |
|  | D | NA | 69.5 | 90.2 | 84.4 | 73.5 | 88.3 |
|  | E | NA | 87.3 | 90.2 | 89.4 | 77.7 | 94.8 |
|  | C + Model | NA | 95.3 | 80.5 | 84.7 | 65.6 | 97.8 |
|  | D + Model | NA | 93.2 | 86.1 | 88.1 | 72.4 | 97.0 |
|  | E + Model | NA | 95.8 | 86.1 | 88.8 | 72.9 | 98.1 |
| Patient-level<br>(Diagnosed with all<br>images) | AI Model | 0.956 | 93.1 | 93.9 | 93.6 | 88.8 | 96.3 |
|  | C | NA | 88.2 | 78.1 | 81.5 | 67.7 | 92.7 |
|  | D | NA | 81.4 | 87.2 | 85.2 | 76.9 | 90.0 |
|  | E | NA | 89.2 | 89.8 | 89.6 | 82.0 | 94.1 |
|  | C + Model | NA | 96.1 | 75.5 | 82.6 | 67.1 | 97.4 |
|  | D + Model | NA | 97.1 | 83.2 | 87.9 | 75.0 | 98.2 |
|  | E + Model | NA | 98.0 | 85.2 | 89.6 | 77.5 | 98.8 |
| Patient-level<br>(Diagnosed with single<br>image) | AI Model | 0.930 | 87.3 | 93.9 | 91.6 | 88.1 | 93.4 |
|  | C | NA | 76.5 | 81.8 | 80.5 | 69.6 | 87.1 |
|  | D | NA | 70.6 | 92.3 | 84.9 | 82.8 | 85.8 |
|  | E | NA | 83.3 | 87.4 | 86.6 | 77.4 | 91.1 |
|  | C + Model | NA | 95.1 | 81.6 | 86.2 | 72.9 | 97.0 |
|  | D + Model | NA | 92.2 | 87.4 | 89.6 | 80.3 | 95.6 |
|  | E + Model | NA | 96.1 | 86.2 | 89.6 | 78.4 | 97.7 |
| Smartphone image | AI Model | 0.902 | 89.2 | 85.7 | 86.9 | 76.5 | 93.9 |
| Video* | AI Model | NA | 94.1 | 94.1 | 94.1 | 94.1 | 94.1 |
|  | C | NA | 100 | 58.8 | 79.4 | 70.8 | 100 |
|  | D | NA | 82.3 | 100 | 91.2 | 100 | 85.0 |
|  | E | NA | 94.1 | 94.1 | 94.1 | 94.1 | 94.1 |

Note\_PPV= Positive predictive value; NPV= Negative predictive value.
\*34 sonographic videos were obtained from 34 infants (17 with BA and 17 without BA).

**Fig. 2.**
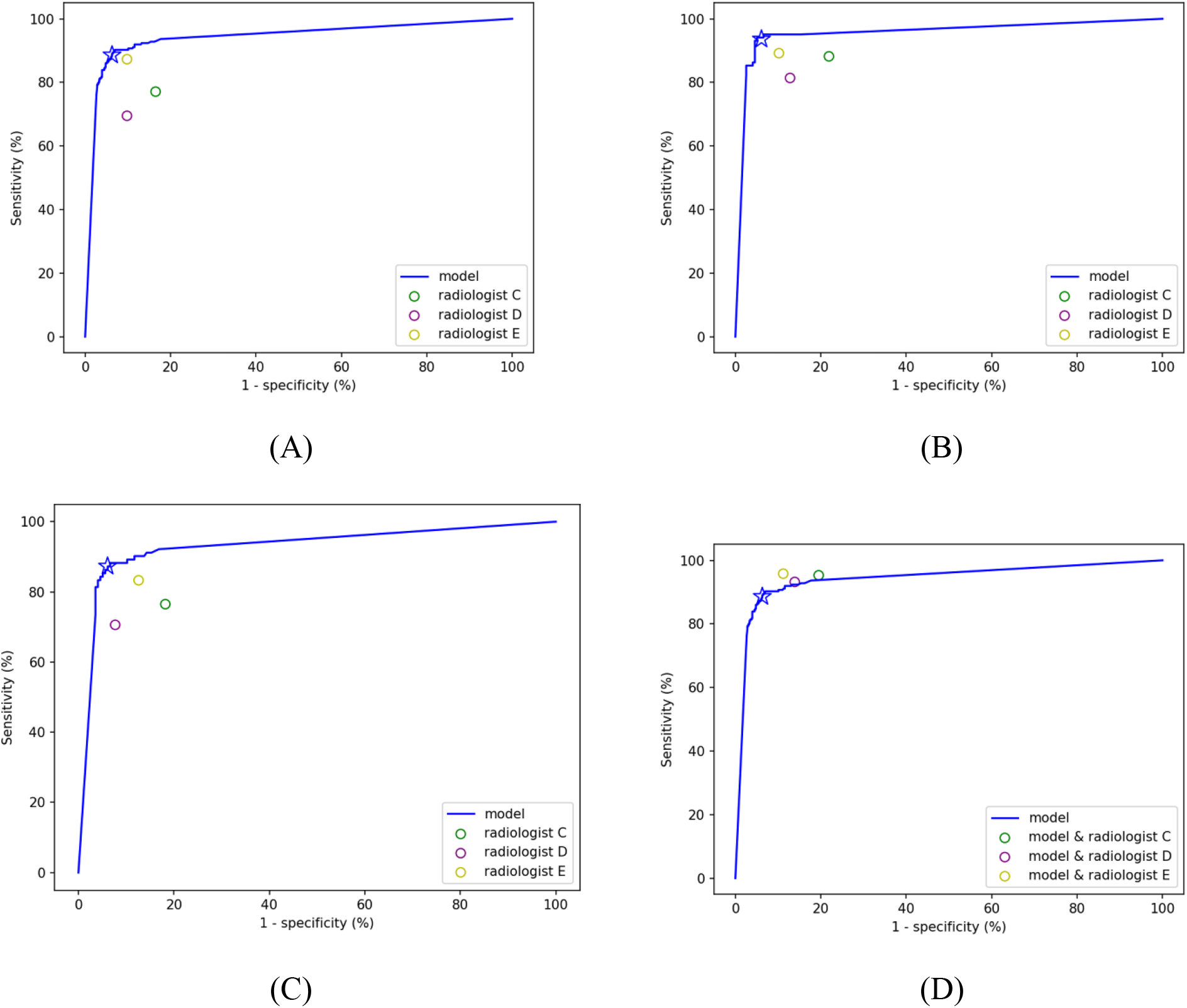
The performance of the ensembled deep learning model, human experts, and the combinations of model and humans for the diagnosis of biliary atresia on the external validation dataset. (A)the ROC curve of the model at image level. (B) the ROC curve of the model at patient level base on majority vote. (C) the ROC curve of the model at patient level based on single image with best image quality for each patient. (D) the performance of the combined deep learning model and human expert (circles) at image level. The circles are above the ROC curve of the deep learning model, suggesting the superior performance of the human-AI combination. The blue star mark represents the performance of the model with the default threshold (0.5) to binarize predictions of the model.

When using the majority vote over the predicted classes of multiple images for each patient, the EDLM achieved an accuracy 93.6%, sensitivity 93.1%, specificity 93.9%, positive predictive value 88.8%, and negative predictive value 96.3% (Table 2, row 8). Another way to obtain patient-level performance is from the diagnosis of a single image for each patient, where the single image was chosen from the multiple images of the patient by a radiologist based on the imaging quality (e.g., less blurry, better view of gallbladder, etc). Such single-image diagnosis by the EDLM achieved an accuracy 91.6%, sensitivity 87.3%, specificity 93.9%, positive predictive value88.1%, and negative predictive value 93.4% (Table 2, row 15). Both the majority vote and the single-image based diagnosis at the patient level by the EDLM outperformed the diagnosis performance of all the three human experts, as seen in Table 2 (rows 9 to 11 and rows 16 to 18) and in the ROC curve (Figure 2B and Figure 2C).

### Combining the diagnosis of theEDLM and expert

Considering the potentially serious consequence of delayed treatment for infants with BA, it is desirable to improve the sensitivity of diagnosis while keeping the specificity at a high level. One possible way to achieve such a goal is to combine the diagnosis of human expert with that of the deep learning model. Here on the external validation dataset, each patient was diagnosed with BA if either the expert or the EDLM thought so. With such combined diagnosis, the sensitivities of three human experts at image level were improved substantially (Expert C, increased from 77.1% to 95.3%; Expert D, increased from 69.5% to 93.2%; Expert E, increased from 87.3% to 95.8%), although the specificities of their diagnosis decreased moderately (Expert C, decreased from 83.5% to 80.5%; Expert D, decreased from 90.2% to 86.1%; Expert E, decreased from 90.2% to 86.1%) (Table 2, rows 5 to 7). Similar findings were found when tested at patient level with multiple images and at patient level with a single image (Table 2, rows 12 to 14 and rows 19 to 21). These findings suggest that the combined approach outperforms not only each expert but also the EDLM, as confirmed from the ROC curve in Figure 2D (also see supplementary Figure S3), particularly in reducing the misdiagnosis of BA.

### Diagnosis based on smartphone photos of sonographic images by the EDLM

In reality, the sonographic machines used for medical examination in hospitals are often not connected to the internet, and it may not be convenient or allowed to extract the original ultrasound images from the machine system. To avoid such obstacle when applying the deep learning model in more medical centers particularly from rural areas, one simple solution is to take a photograph of the sonographic image by a smartphone and then send the photo to a remotely located AI system for intelligent diagnosis. However, the image quality of the photograph would be inevitably affected by this imaging process, e.g., with more noise included or shape and texture of gallbladder regions deformed (Figures 3A Right, 3B Right). Therefore, it would be desirable if the deep learning model can still work well when applied to the analysis of such smartphone photos.

**Fig. 3.**
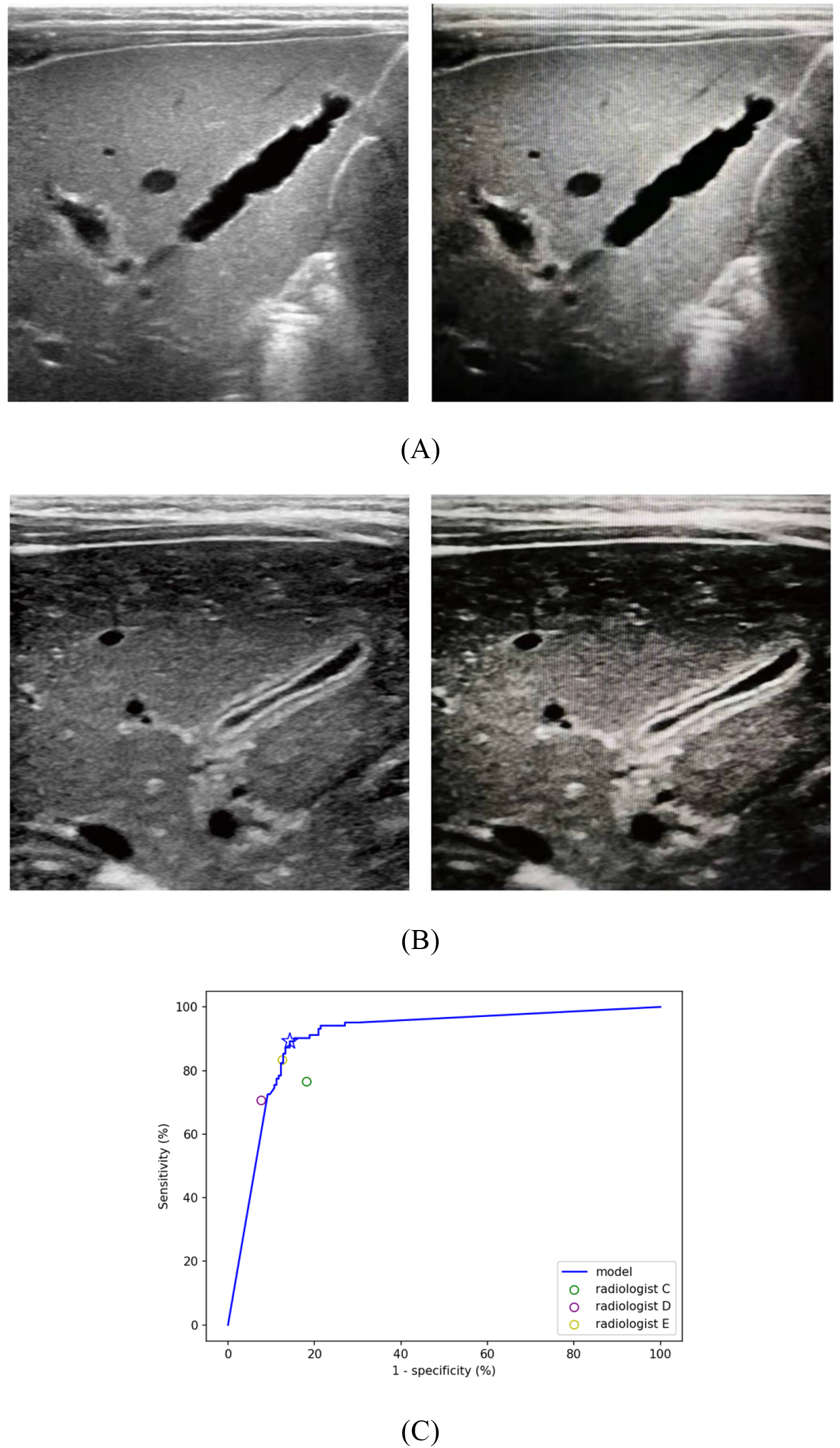
Diagnosis based on smartphone photos of sonographic images by the deep learning model. (A) an exemplar original image from a patient with BA (Left) and the smartphone photo of the image (Right). (B) an exemplar original image from a patient without BA (Left) and the smartphone photo of the image (Right). (C) the ROC curve of the model for the diagnosis of BA on the smartphone images of external validation dataset, with three human experts’ performance on the original clean external validation dataset for comparison.

To evaluate the robustness of the EDLM in this case, one original image per patient (as mentioned above for the single-image diagnosis) from the external validation dataset was pictured by a smartphone (HUAWEI P10, Rear Camera: 12 million pixels) (Figures 3A, 3B), with the original image information kept as much as possible during picturing (e.g., by making camera viewing direction perpendicular to the machine screen). Smartphone photos were saved in the JPEG format. As done for the original images, the region of gallbladder was extracted from each photograph and then fed into the EDLM for intelligent diagnosis. Although the EDLM was trained with the original clean images of the training cohort, the diagnosis of the smartphone images by the model resulted in an accuracy 86.9%, sensitivity 89.2%, and specificity 85.7% (Table 2, row 22). The AUC value (0.902) is slightly lower than that tested with the original images (AUC=0.930), but the ROC curve (Figure 3C) together with the prediction performance (Table 2, row 22) suggests that such diagnosis is still comparable to all three human experts who diagnosed based on the original *clean* images.

Considering the promising external validation result based on smartphone photos, a smartphone app has been developed and released (Figure 4), from which users can freely upload photos of ultrasound images and interactively locate the gallbladder regions. The software would send photos to and collect prediction results from a cloud platform running the EDLM. An initial prospective study(sonographic gallbladder images were from multicenters photographed by different radiologists) with 71 BA patients and 103 non-BA patients (one photo per patient) showed that the app performed similarly well, with an accuracy 85.6%, sensitivity 85.9%, specificity 85.4%, and AUC value (0.856). The small variation in performance between the prospective study and the above external validation is probably due to the uncontrolled picturing conditions in the prospective study, where different users may use different smartphones in varying lighting environments. Such smartphone app provides the opportunity to help clinicians improve their diagnosis performance particularly for hospitalsin rural area.

**Fig.4.**
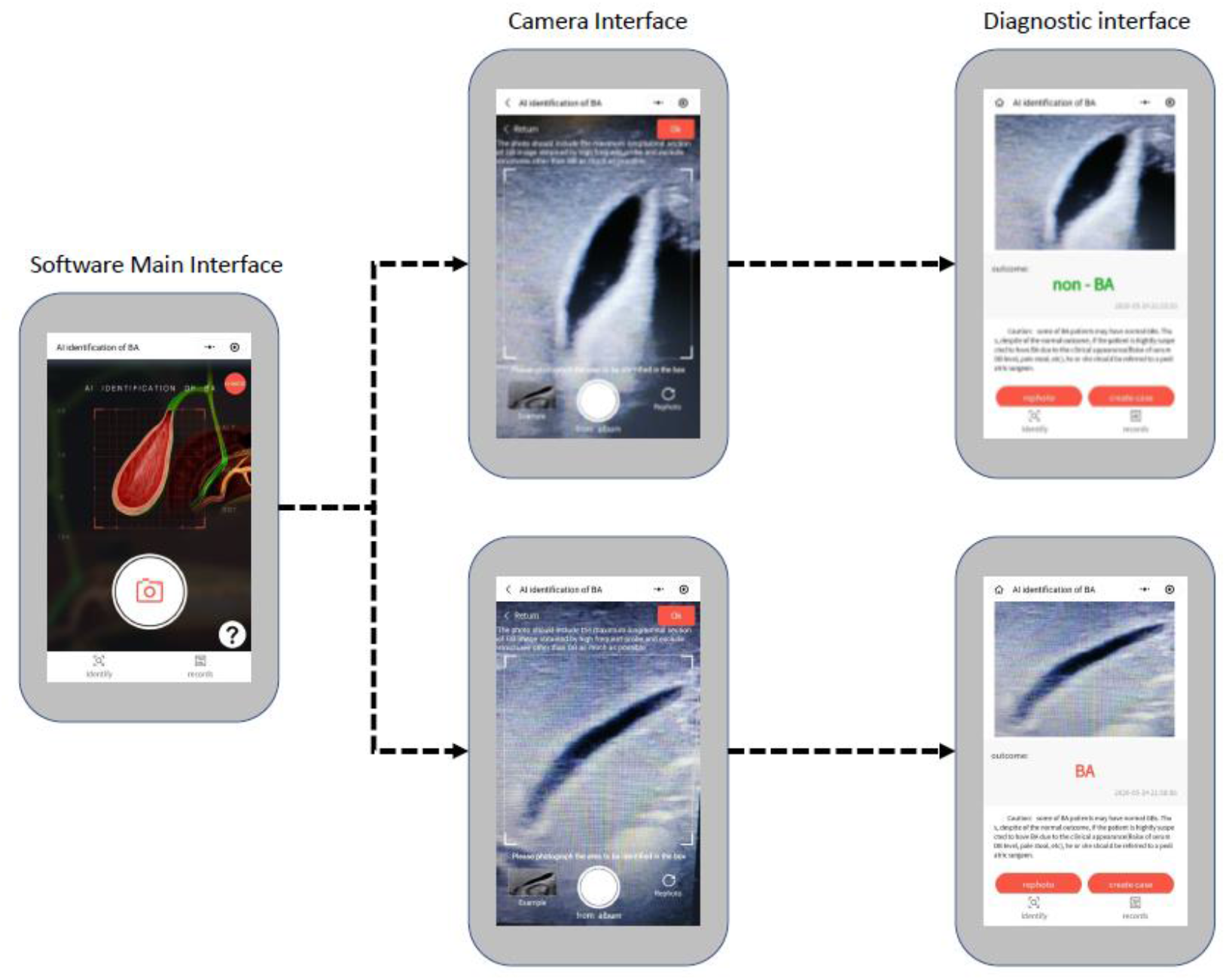
The interface for each step of the smartphone app based on the ensembled deep learning model.

### Diagnosis based on sonographic videos by the EDLM

In practice, human radiologists make diagnosis not based on observing one or a few static sonographic images but by dynamically observing the gallbladder region with real time US scanning. Also, it would be inconvenient for radiologists to select one or a few static images and then draw bounding boxes surrounding the gallbladder before sending the images to the intelligent diagnosis system. Therefore, it would be ideal if the intelligent diagnosis system can make fully automatic diagnosis just based on the recorded video sequence of sonographic images. To achieve this goal, we trained an auto segmentation model(see supplementary materials) and an initial prospective study was performed with a collection of 34 sonographic videos obtained from 34 infants (17 with BA and 17 without BA). The diagnostic performance of EDLM was compared with performance of three human experts, each of whom independently made diagnosis by reviewing videos and were blinded to other clinical information.

Based on the fully automatic diagnosis process (figure 5), 16 out of the 17 BA videos were correctly diagnosed containing BA (sensitivity 94.1%), and 16 out of the 17 non-BA or healthy videos were correctly diagnosed containing no BA (specificity 94.1%). Compared to the diagnosis performance from the experts (Table 2, last 3 rows), the EDLM is comparable to three experts. More evaluations showed that the diagnosis performance of the EDLM changes little when varying the model hyperparameters, such as changing the percent of the selected images from 20% to 10% and changing the percent of diagnosed BA images from 10% to 30%, suggesting strong robustness of theEDLM.

**Fig.5.**
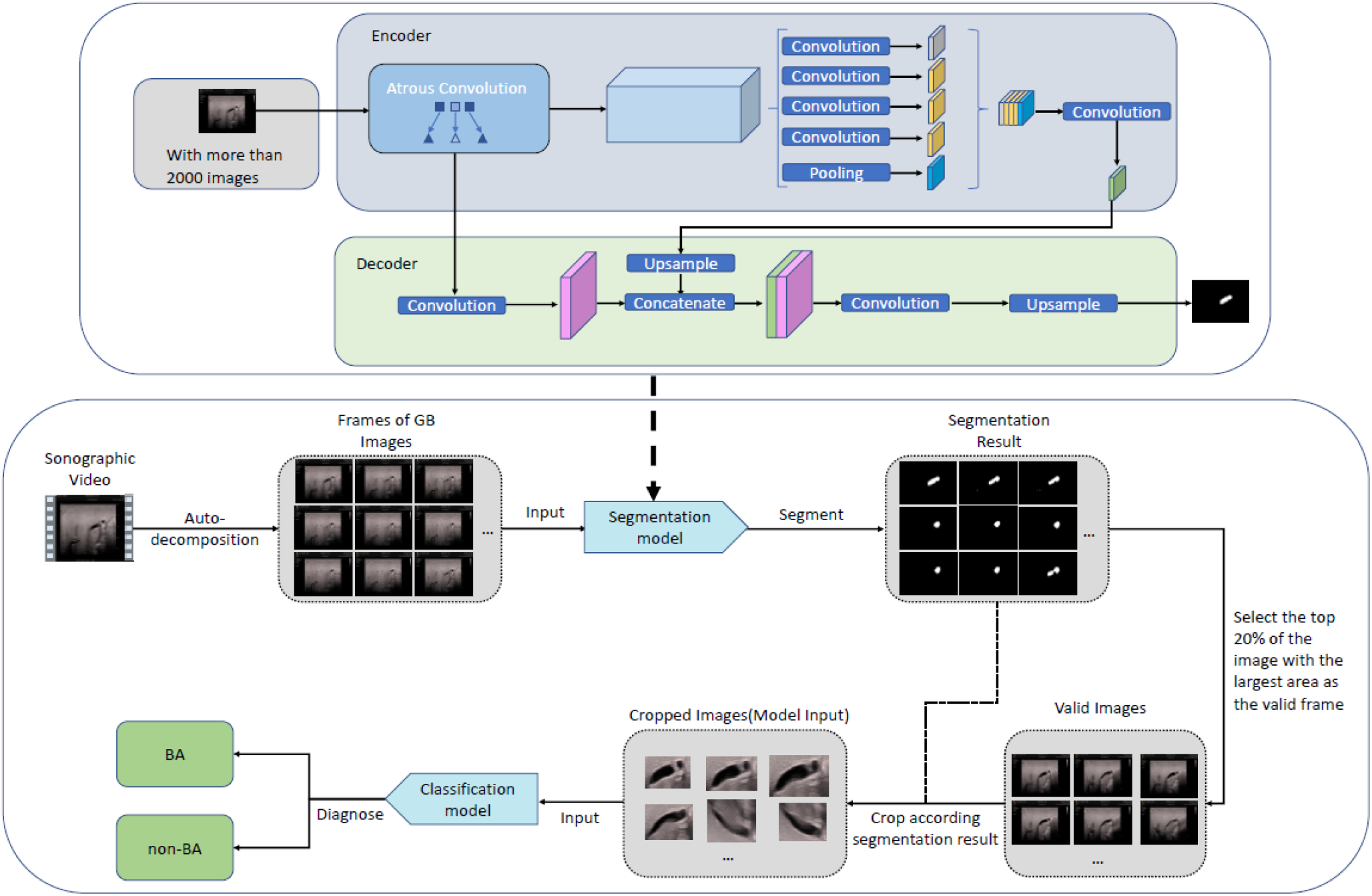
The diagnostic process for each sonographic gallbladder video.

### Initial attempt to interpret AI diagnosis

One widely-used method to explore black-box of AI diagnosis is the class activation map (CAM) which can provide the attended image region(s) for each specific prediction from the model ^30^. Based on the attended region from CAM, people may infer why the model makes the current prediction for each image (e.g., “because the model focused on the gallbladder region and therefore used the visual features within this region to make the decision”). If the attended region obtained by CAM covers or partly covers the regions used by human experts for diagnosis (“Consistent” in supplementary Table S3), it may improve the sense of trust in the AI model for the current diagnosis. Otherwise, if the attended region obtained by CAM does not cover any region of interest used by experts (“Inconsistent” in supplementary Table S3), this may indicate that the AI model does not use appropriate visual features to make current (either correct or incorrect) decision. Of all the external validation images, detailed inspection showed that 98.1% were consistent in decision making between the EDLM and human experts. Of the correctly diagnosed external validation images, 97.9% were consistent (Supplementary Table S3, row 3; also see Figure 6A) and 2.1% were inconsistent (Supplementary Table S3, row 3; Figure 6B); Of the incorrectly diagnosed external validation images, 100% were consistent (Supplementary Table S3, row 6; Figure 6C).

**Fig.6.**
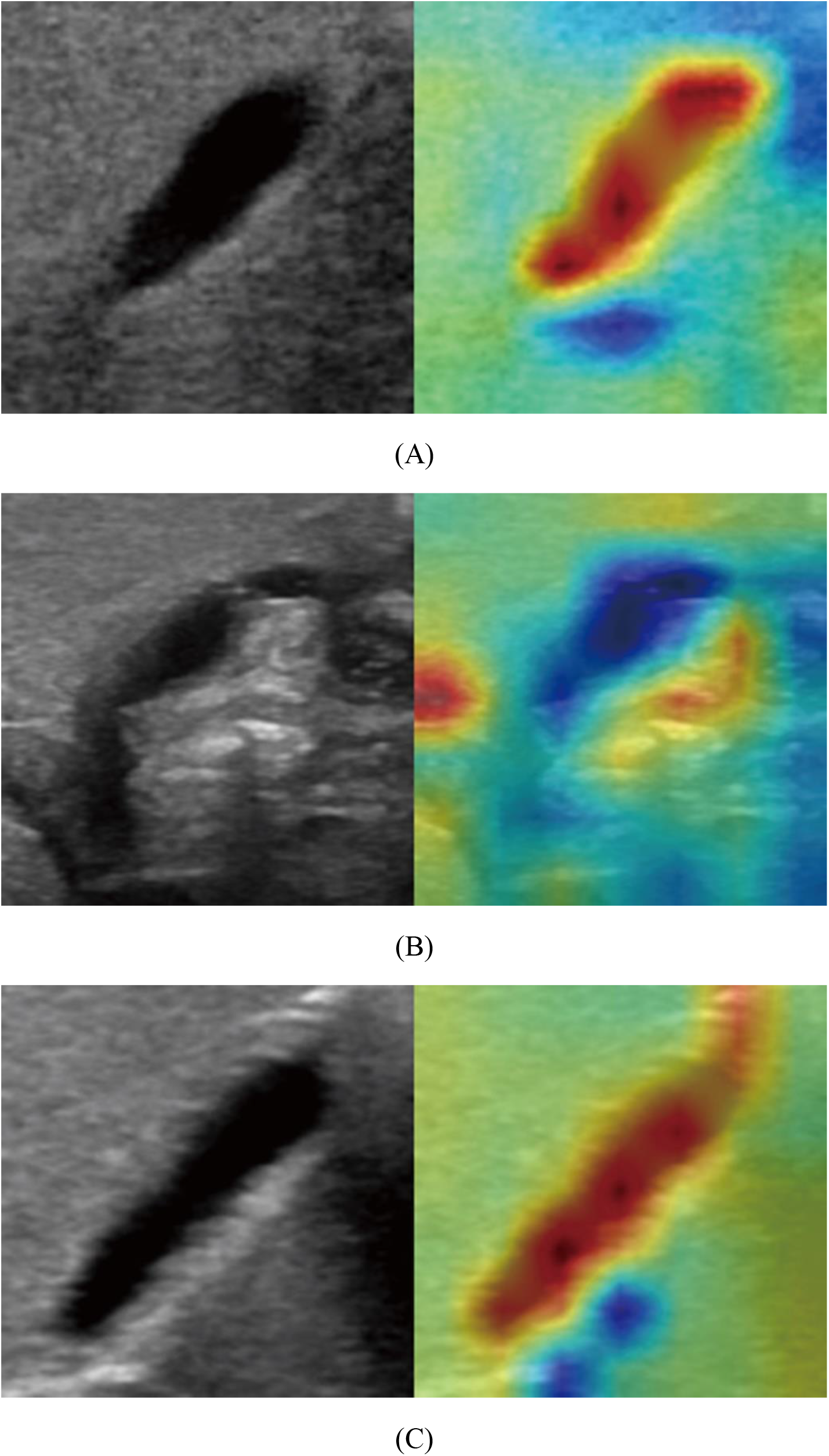
The attended regions during diagnosis by the ensembled deep learning model (with reddish regions corresponding to more attention in the heatmap on each row). (A) the image diagnosed correctly by the model, and the region of interest was consistent between the model and human expert. (B) the image diagnosed correctly by the model, and the region of interest was inconsistent between the model and human expert. (C) the image diagnosed incorrectly by the model, and the region of interest was consistent between the model and human expert.

## Discussion

In this multicenter study, we trained and validated a state-of-the-art EDLM based on sonographic gallbladder images for the diagnosis of BA. The EDLM outperformed human experts on both internal training and external validation cohorts. Moreover, combined with the prediction of the EDLM, the sensitivities of human experts in identifying patients with BA were substantially improved from 69.5%∼87.3% to 93.2%∼95.8% in image-level diagnosis, and were even better than that of the EDLM alone. A higher sensitivity would lead to fewer misdiagnosis and hence benefit patients with suspected BA in clinical practice. Hence, all these findings indicate that the EDLM could not only be used to help diagnose BA in primary hospitals lacking experts, but also help experienced experts to further improve their performance in the diagnosis of BA.

We also evaluated the EDLM with gallbladder photos taken by a smartphone. Although the image quality of smartphone photos was inevitably downgraded compared to the original clean images, surprisingly, the model still performed well, with similar accuracy but higher sensitivity than the experts. Another prospective study with the developed and released smartphone app showed similar diagnosis performance. This opens an opportunity of remote and convenient online diagnosis especially for rural and underdeveloped regions without experts. In China, sonographic machines for medical diagnosis are usually not allowed to connect to the internet. However, the expert-level performance of the EDLM on the smartphone app, together with the nation-wide mobile networks and low-price smartphones, would make it easy and convenient for clinical staff even in remote underdeveloped areas to upload gallbladder photos with smartphones for online and real-time diagnosis consultancy. Such photo-based online consultancy would largely improve the diagnosis accuracy particularly for those radiologists with less experience or from underdeveloped regions.

In addition, an initial video-based intelligent diagnosis showed that theEDLM, together with automatic selection of relevant images and localization of gallbladder regions, can more accurately diagnose BA than human experts. Such video-based diagnosis avoids the manual effort in image selection and gallbladder region localization by radiologists, and could be potentially embedded into the existing diagnostic ultrasound system for fully automated diagnosis of BA during medical examination.

The initial attempt to interpret the model’s predictions showed that the model also attended to the gallbladder regions during diagnosis as human experts did. However, among a small proportion of the correct diagnoses, the model made decisions just based on the visual features outside the gallbladder regions. There were 2.1% of inconsistency among correctly diagnosed external validation images, which indicates the model can make a correct diagnosis by recognizing features other than gallbladders in some circumstances. This suggests that there might exist certain non-gallbladder features associated with BA. More investigation is necessary to explore the potentially novel biomarkers for the diagnosis of BA.

Deep learning models are usually powered by a large scale of dataset ^31^. However, BA is a rare disease with low incidence, making it challenging to obtain large dataset as for other diseases ^26, 27, 28, 29^. To alleviate the potential over-fitting issue due to limited training dataset, we applied a few number of effective strategies for model training, including the ensemble learning, data augmentation, class weight for the imbalanced dataset between the BA and the non-BA classes, dropout of neurons during learning, and transfer learning from a pre-trained deep learning model based on large-scale natural images. Experiments showed that these strategies largely improved the generalizability of the deep learning model particularly when evaluated on the external validation dataset, suggesting that such strategies may be adopted in prospective studies relevant to medical image classification.

In most of this study, the gallbladder region in each image need to be manually located with a form of bounding box provided by radiologists, which inevitably would increase burden on human experts during diagnosis. This issue can be avoided by automatically detecting the region of gallbladder from each image, which is feasible based on the recently developed deep learning models like Faster R-CNN ^32^ and will be part of the future work. Furthermore, the initial investigation of model interpretation told us, if there were vascular or intestinal gas interference around the gallbladder, the model might mistakenly identify these interfering tissues as gallbladder and made a diagnosis partly based these non-gallbladder regions. Automatic precise localization of gallbladder could make the AI model focus on the correct gallbladder region and therefore potentially further improve the performance of intelligent diagnosis. This may be achieved by the recently developed deep learning based semantic image segmentation models like the U-Net ^33^ and DeepLab ^34^. The more automatic precise localization of gallbladder regions would also enable more accurate video-based intelligent diagnosis. In addition, recent study ^35^ showed that the AI performance could be improved when using three-dimensional sonographic data. Therefore, one possible future work is to use sonographic volume data to potentially further improve the performance of the deep learning model.

In conclusion, we developed an EDLM that outperforms human experts in diagnosis of BA based on a relatively small-scale sonographic gallbladder images acquired from five different hospitals. The generalization capability of the model was confirmed with an external validation dataset obtained from another six hospitals. Moreover, this model is potentially deployable in multiple application scenarios, such as remote diagnosis based on a smartphone app to conveniently help the unexperienced radiologists in primary hospitals, diagnosis based on the combined predictions from the model and human radiologists to further improve the diagnosis sensitivity even for experienced radiologists in tertiary hospitals. To the best of our knowledge, this is the first deep learning model for the diagnosis of BA based on sonographic gallbladder images. Since there are still lots of underdeveloped regions lacking sufficient healthcare support and experts for diagnosing BA all over the world, the application of the EDLM in clinical practice would benefit those jaundiced infants with suspected BA.

## Materials and Methods

### Patients and data collection

This multicenter study was approved by the institutional Clinical Research Ethics Committee of the First Affiliated Hospital of Sun Yat-sen University, and written informed parental consent was obtained before collecting the sonographic images from each patient. Prospective research of this study was also registered at www.chictr.org.cn (ChiCTR1800017428).

Infants younger than 5 months old with hyperbilirubinemia (serum direct bilirubin level >17.1 umol/L and the ratio of direct to total bilirubin level >20%) ^36^ and suspected of BA were initially selected from 11 hospitals between January 2010 and June 2019 (Figure 7). The exclusion criteria for patients are as follows: (1) the final diagnosis was unclear; (2) jaundice was caused by bile duct obstruction to which abdominal mass compression gave rise; (3) the patient had a history of abdominal surgery; (4) the visualization of gallbladder was indeterminate. For the patients satisfying (4), the diagnosis is highly suggestive of biliary atresia and referral to the experienced center for further examination would be recommended. In order to expand the sample size, we also randomly selected some infants from the same 11 hospitals who did not have any known liver diseases and were considered as non-BA with a normal transcutaneous bilirubinometer test. Finally, a total of 1100 patients with suspected BA and 339 infants without jaundice were enrolled. Of the 1100 patients with suspected BA, 432 infants had BA and 668 infants had non-BA. All diagnoses were confirmed by intraoperative cholangiography under laparoscopy, percutaneous ultrasound-guided cholecystocholangiography, liver biopsy or follow up.

**Fig.7.**
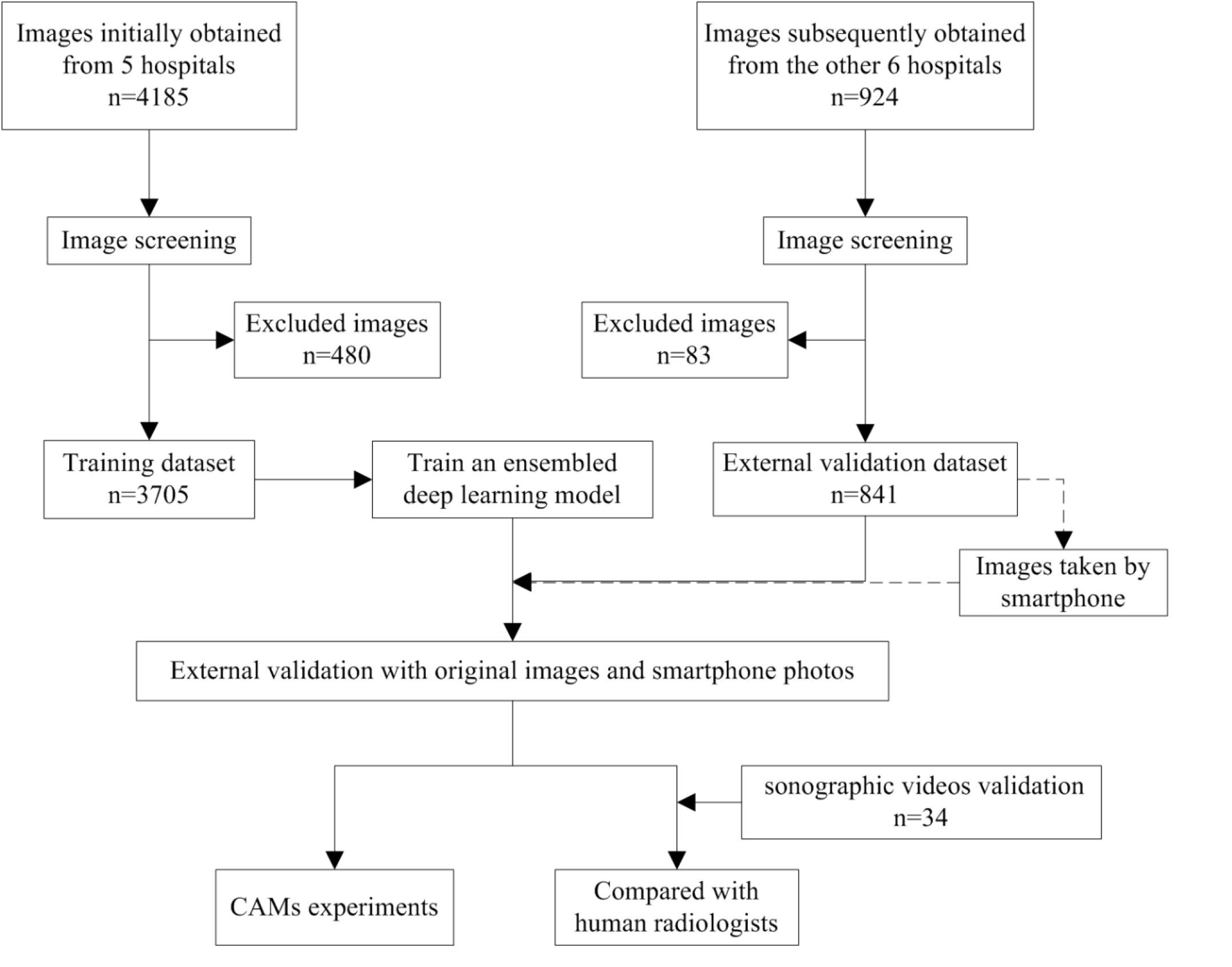
Flow chart of the study.

For each infant, sonographic gallbladder images were acquired either prospectively or retrospectively. The prospective image acquisition need to satisfy the following criteria: (1) images were acquired after the patient fasted for at least 2 hours; (2) gallbladder was detected by high-frequency transducers (> 7 MHz); (3) a complete outline of the gallbladder long axis was included; (4) there was no mark or caliper within the image; (5) the depth of the image is less than 5 cm; (6) the image resolution is large enough (often larger than 300-by-300 pixels); (7) at least 2 independent gallbladder images were obtained from each patient. When images were acquired retrospectively, at least the criteria (1), (2), (3), (4) and (6) need to be satisfied.

All images that were potentially available were reviewed by a senior sonography expert (L.Y.Z.) and those with poor quality were excluded. We finally retrospectively and prospectively obtained 3705 sonographic gallbladder images (925 from 330 patients with BA, 2780 from 811 patients without BA) from the principal hospital and 4 collaborating hospitals as training cohort, and prospectively obtained 841 sonographic gallbladder images (236 images from 102 patients with BA, and the other 605 images from 196 patients without BA) from the remaining 6 collaborating hospitals as external validation cohort (Supplementary Table S4). Considering that each image includes irrelevant regions (e.g., dark regions close to image boundaries and text information around the top regions), a bounding box containing the entire gallbladder was manually drawn with the free software ImageJ (version 1.52a) by two radiologists (W.Y.Z. and W.H.G.), and then a senior doctor (L.Y.Z.) double checked and ensured the bounding box was selected appropriately.

### Diagnosis by Human Experts

In order to evaluate the efficacy of the deep learning approach, the performance of human experts was provided in advance for direct comparison between the AI model and humans. To obtain the patient-level diagnosis performance, each random-ordered patient’s image data in the training cohort was presented and diagnosed as either BA or non-BA independently by two human experts (J .X.L. and C.Y.), and each patient’s image data in the external validation cohort was presented and diagnosed independently by the other three human experts (Z.J.W., D.C. and X.X.D.), both only based on all the available (often 1-3) images for each patient diagnosis. All five experts had more than 10 years of experience with pediatric ultrasound. Similarly, to obtain the image-level diagnosis performance, each image without any patient ID information was presented randomly and diagnosed independently by the same experts as for the patient-level diagnosis. All these five experts have not read any of the patient images before attending this study and had no access to any other patient information (e.g., clinical history, other imaging results, etc.) during their diagnosis.

### Ensembled deep learning framework

In this study, two types of effective AI techniques called deep convolutional neural networks (CNNs) and ensemble learning were adopted and combined together for intelligent diagnosis of BA. Multiple (e.g., 5 here) CNNs were trained with the training cohort and then the output predictions of these CNNs were averaged to predict the class label of each image in the validation dataset, resulting in an EDLM (Figure 8A). Specifically, the training cohort was randomly separated into five complementary subsets (i.e., 5-folds), each containing the images of an equivalent number of patients. Then, each CNN was trained with four subsets and the training was stopped when the performance of the CNN started to decrease on the remaining subset. The subset used to determine time point to stop the training of each CNN was unique (e.g., subset 5 for first CNN, and subset 1 for second CNN), which also means that the combination of four subsets for training each CNN was also unique (e.g., subset 1-4 for first CNN, and subsets 2-5 for second CNN). In this way, we not only solved the issue about when to stop training a CNN, but also make the five trained CNNs a bit more diverse from each other, where the diversity among CNNs would improve the generalization ability of the ensembled model as confirmed in the empirical evaluation. The adopted CNN model Se-ResNet (Supplementary Figure S4) and the training of each Se-ResNet (Figure 8B) were described in detail in the supplementary material.

**Fig.8.**
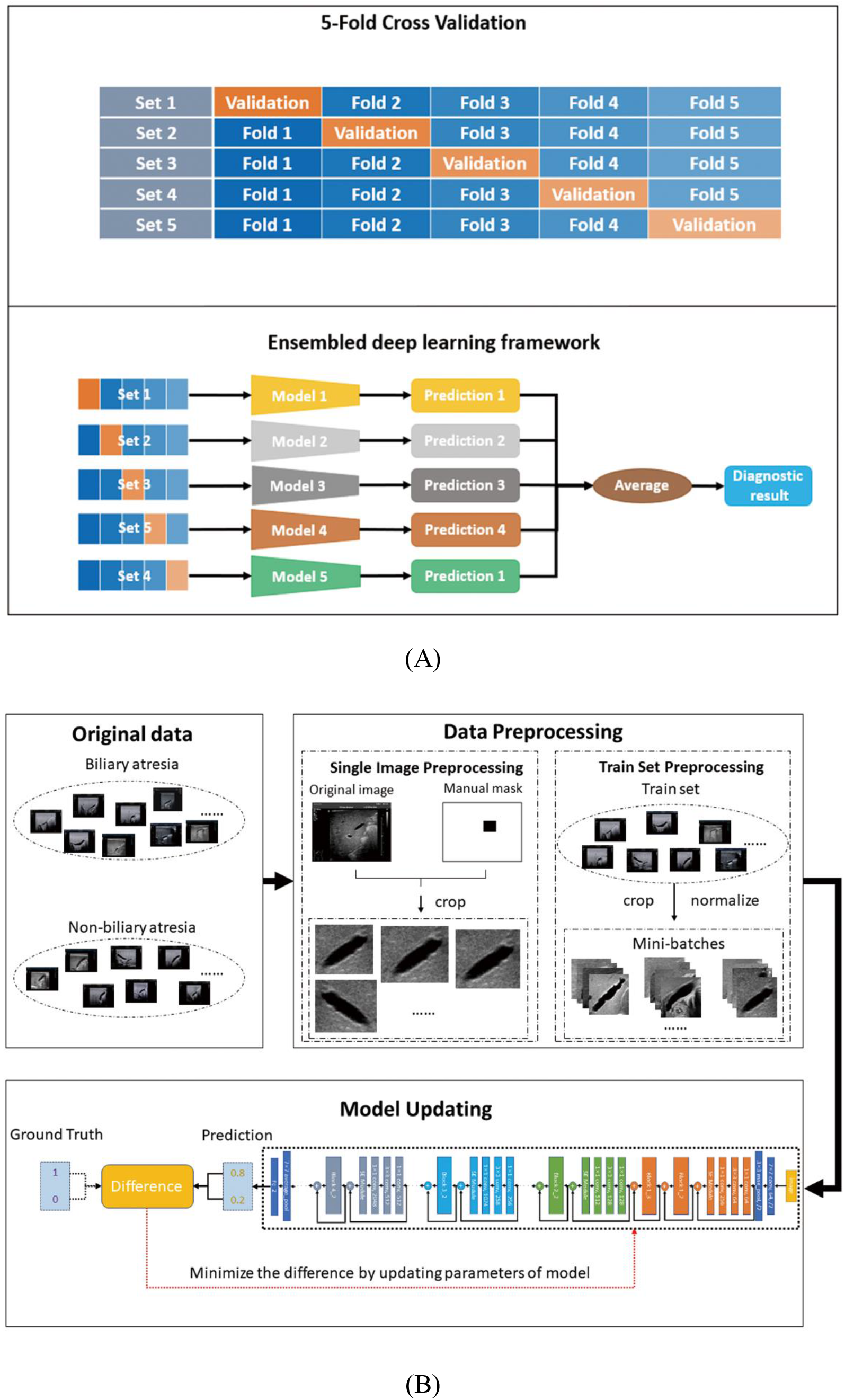
The ensembled deep learning approach for this study. (A) the ensembled deep learning framework. (B) the training process for each individual CNN model.

### Measurements of the diagnostic performance

The performance of each EDLM was evaluated on the test (validation) dataset, with the test dataset varied for different purposes (as seen in the Results section). By comparing the predicted classes from the model with the ground-truth classes obtained in advance over all the test images, the sensitivity, specificity, accuracy, positive predictive value (PPV), negative predictive value (NPV), and the area under the ROC curve (AUC) of the ensembled model were calculated. At the image level, the ROC curve of the EDLM was generated by varying the threshold for the output prediction of the model, where the threshold was used to binarize the model’s continuous output. Different thresholds could lead to different binary predictions of the model for each image and therefore result in different sensitivities and specificities on the test dataset. Similarly at the patient level, a specific threshold would lead to the specific binary predictions for the (often multiple) images of each patient, and therefore result in one specific binary prediction for each patient after the majority voting over the multiple binary predictions of the images from the same patient. Then, by varying the thresholds, one ROC curve would be generated based on the sequence of sensitivities and specificities at the patient level. In addition, for comparison, the above measures except the AUC were also obtained for each human expert based on their diagnosis results and the ground-truth classes for the test images.

## Data Availability

Data referred to in the manuscript is available if required to the corresponding author.

## Acknowledgement

We thank Prof. Yunchao Chen from Xiang’an Hospital of Xiamen University and Dr. Qi Yu from Sanya City Womenfolk and Infant Health Care Hospital for providing data to support this study.

## Author contributions

**W**.**Y**.**Z**. and **Y**.**Y**., images processing, experiments performing, data analysis, drafting; **C**.**Y**., **J**.**X**.**L**., **X**.**X**.**D**., **Z**.**J**.**W**. and **D**.**C**., images providing and images diagnosis; **Q**.**H**.**L**., **F**.**Q**., **J**.**J**.**Z**., **H**.**J**., images providing; **Z**.**H**.**L**., manuscript revising; **W**.**H**.**G**.; image processing; **X**.**Y**.**X, R**.**X**.**W**. and **L**.**Y**.**Z**., study design, experiments performing, data analysis, drafting, accountability for all aspects of the work. All authors have seen and approved the final version of the manuscript.

## Abbreviations

BA: biliary atresia
EDLM: ensembled deep learning model
KPE: Kasai portoenterostomy
AI: artificial intelligence
CNNs: convolutional neural networks
AUC: area under receiver operating characteristic curve
CAM: class activation map

